## Supplementary Figures for "Cross-tissue meta-analysis of blood and brain epigenome-wide association studies in Alzheimer’s disease"

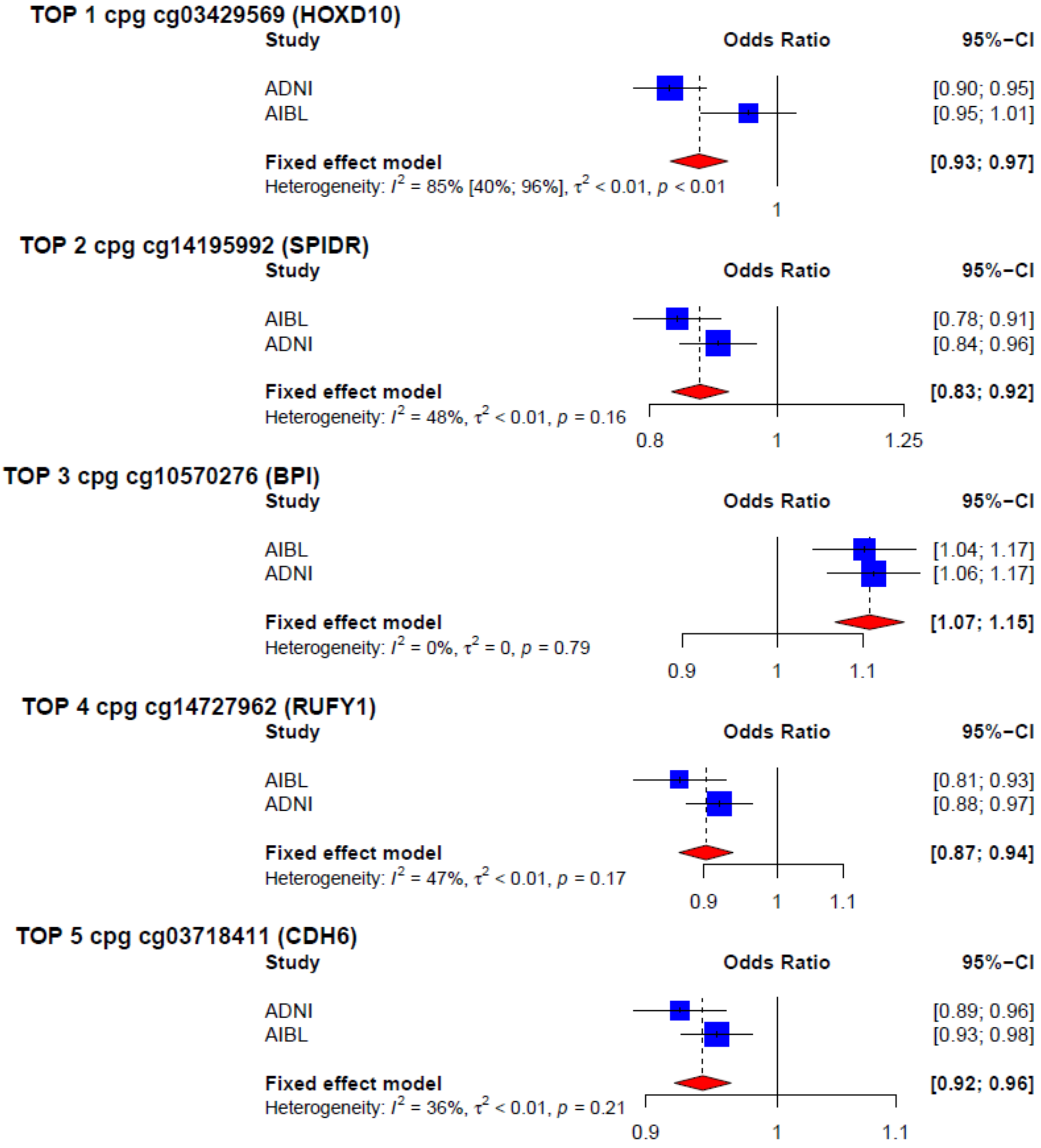


**Supplementary Figure 1** Forest plots for the 5 FDR-significant CpGs (i.e., FDR < 0.05) in meta-analysis of ADNI and AIBL blood samples datasets. Shown are odds ratios and confidence intervals that describe changes in odds of AD (on the multiplicative scale) associated with a one percent increase in DNA methylation beta values (i.e., increase in beta values by 0.01) after adjusting for covariate variables.

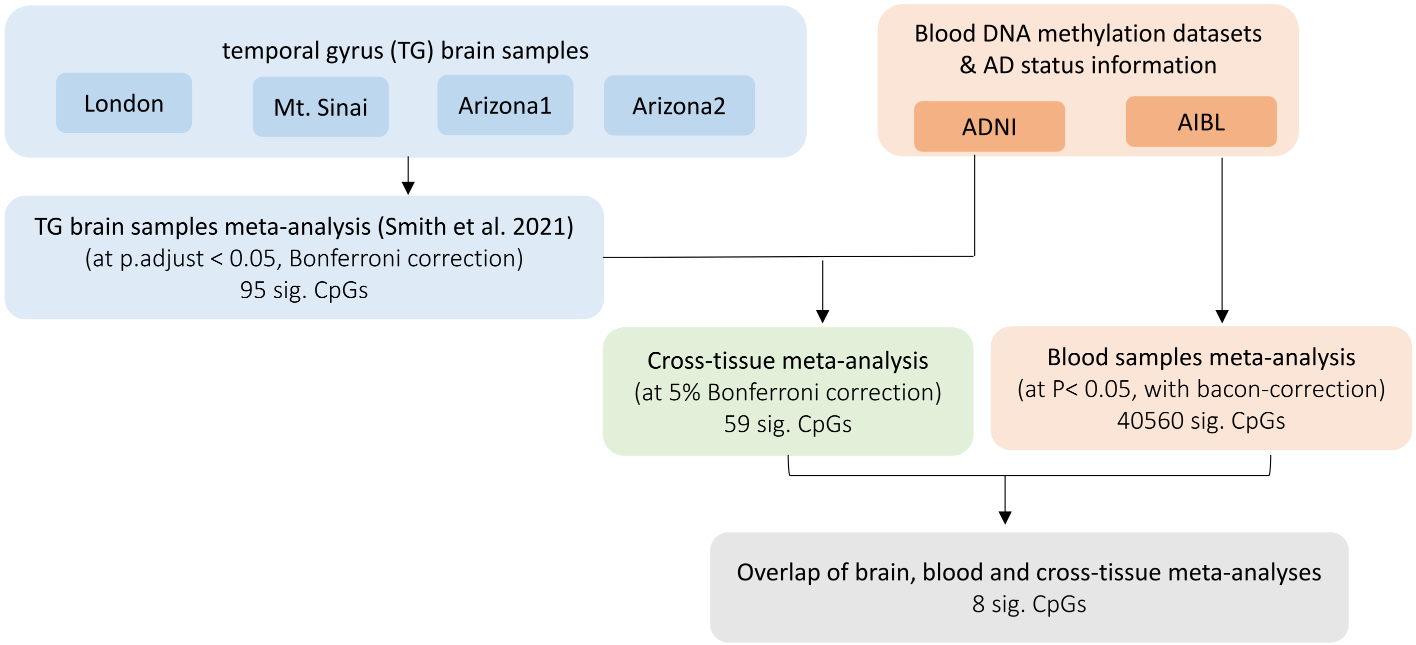


**Supplementary Fig 2** Workflow for prioritizing differentially methylated CpGs associated with AD pathology (in temporal gyrus brain samples) and AD diagnosis (in blood samples). Genomic corrections were performed using the bacon method (PMID: 28129774) in the analysis of all individual datasets. The *P*-values for significant CpGs associated with AD Braak stage in the temporal gyrus region were obtained from Supplementary Table 3 of Smith et al. (2021) (PMID: 34112773).

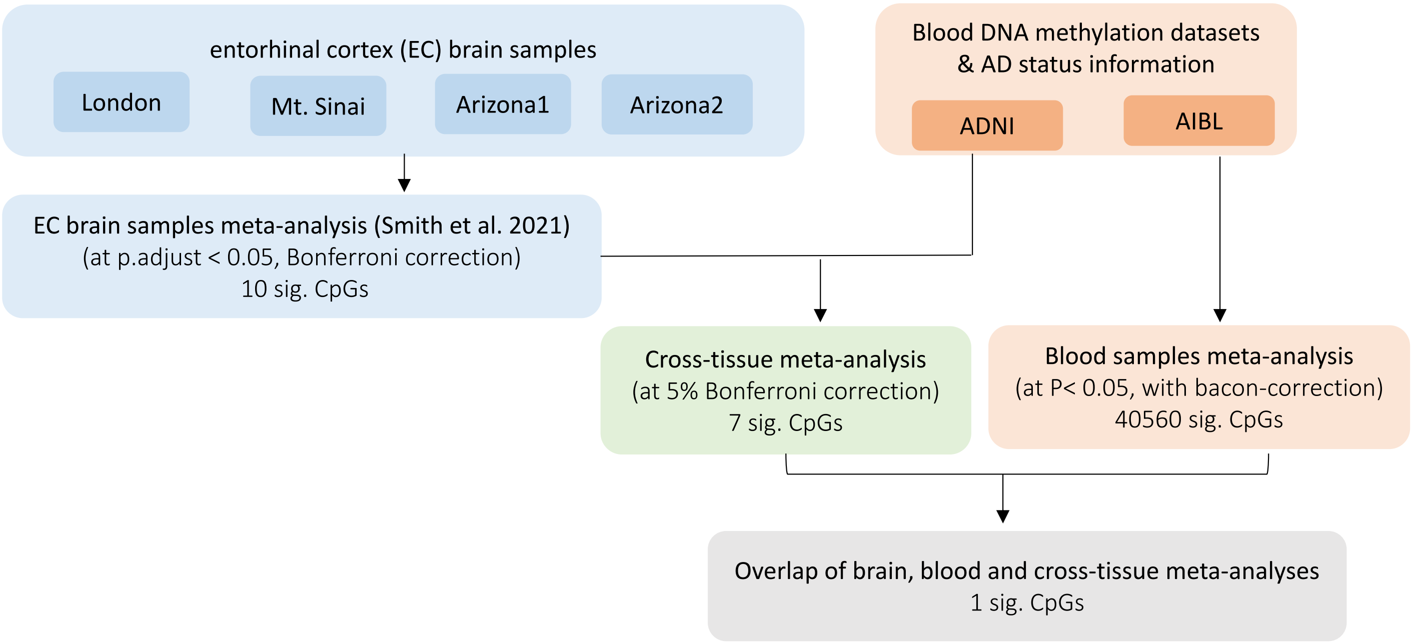


**Supplementary Fig 3** Workflow for prioritizing differentially methylated CpGs associated with AD pathology (in entorhinal cortex brain samples) and AD diagnosis (in blood samples). Genomic corrections were performed using the bacon method (PMID: 28129774) in the analysis of all individual datasets. The *P*-values for significant CpGs associated with AD Braak stage in the entorhinal cortex region were obtained from Supplementary Table 5 of Smith et al. (2021) (PMID: 34112773).

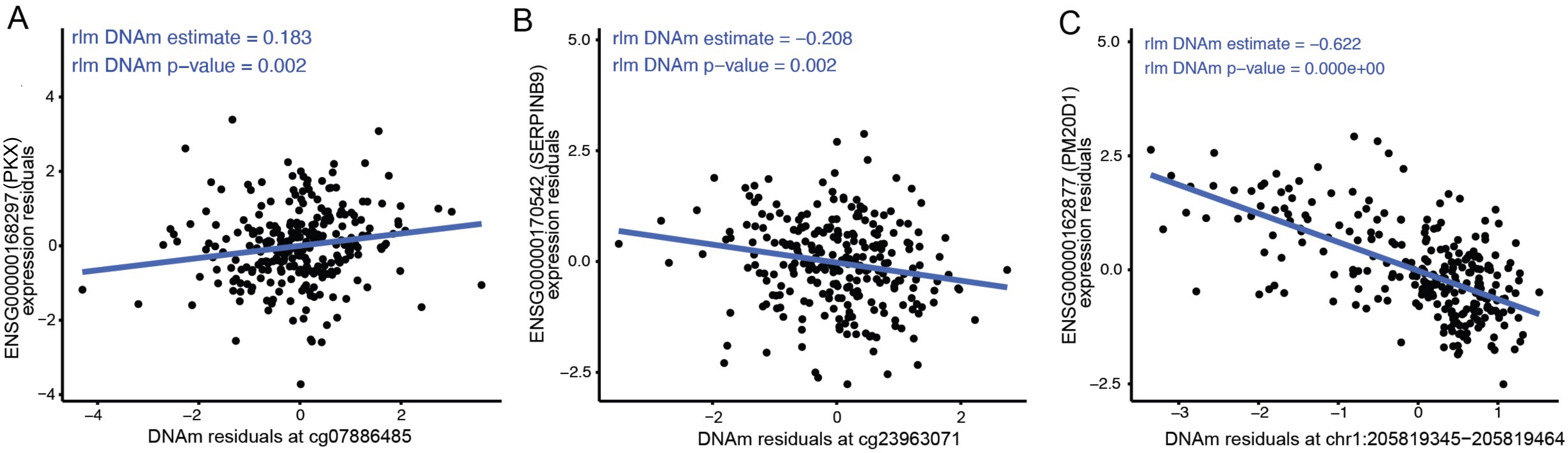


**Supplementary Figure 4** Among the 50 CpGs that reached P < $10^{-5}$ and 9 DMRs with 5% Sidak corrected P-values in AD vs. CN blood samples meta-analysis, 3 CpG-gene or DMR-gene pairs reached 5% FDR significance in the analysis of ADNI blood samples with matched DNAm-RNA data. **A** cg07886485 – PXK, **B** cg23963071-SERPINB9, **C** chr1:205819345-205819464-PM20D1

**A**

| | |
|---|---|
| Genome of reference | hg19 |
| Region ID | chr1:244486252−244486253 |
| Probe ID | cg16908123 |
| Target gene ID | ENSG00000173728 |
| Target gene Symbol | C1orf100 |
| TF gene ID | ENSG00000020633 |
| TF gene Symbol | RUNX3 |
| TF role | Repressor |
| DNAm effect | Attenuating |

| Target ~ TF + DNAm Quant. Group + TF * DNAm Quant. Group | Estimate | P−Values |
|---|---|---|
| Direct effect of DNAm | 0.205 | 0.187287 |
| Direct effect of TF | −0.541 | 4.78e−06 |
| Synergistic effect of DNAm and TF | 0.762 | 6.58e−06 |

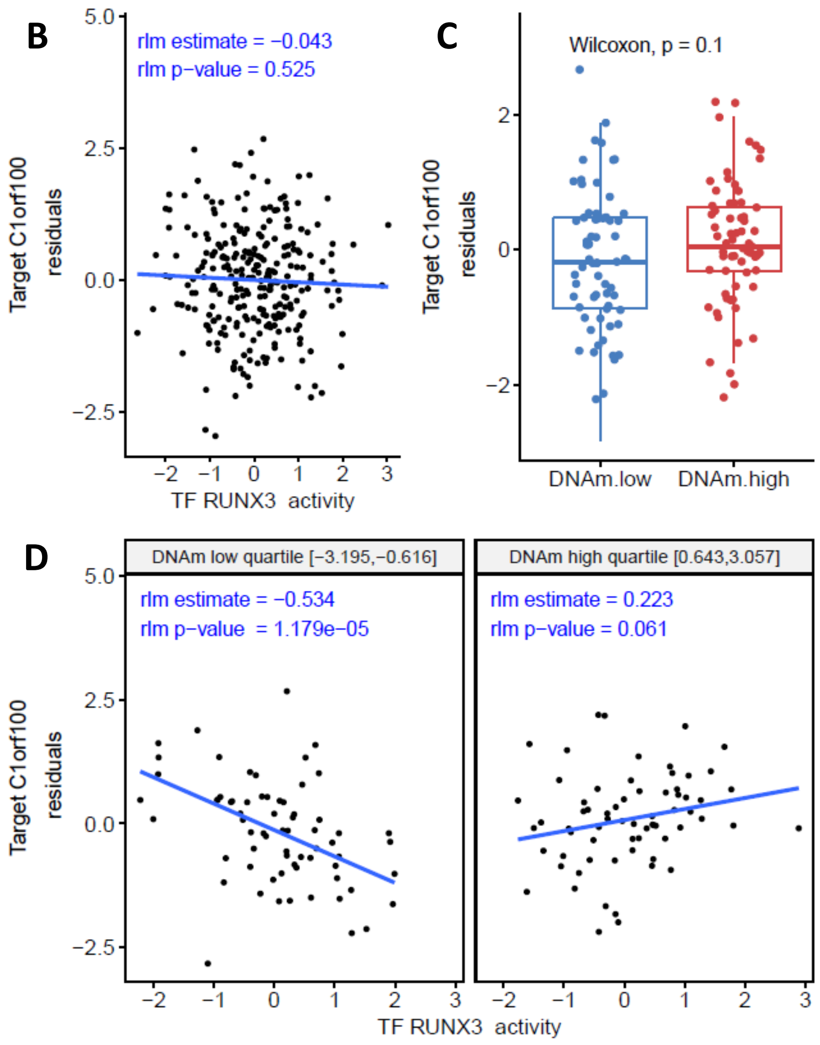


**Supplementary Figure 5** Results from MethReg analysis of ADNI blood samples showed in the cg16908123-RUNX3-C1orf100 triplet, DNAm attenuated TF activity (P-value for DNAm × TF interaction = $6.58 \times 10^{-6}$). **A** Meta data for the CpG-TF-target gene triplet and results of fitting robust linear model to the triplet dataset. **B** When all samples are considered, no significant TF-target gene association was observed. **C** Comparison of target gene expression between the DNAm groups showed no association between DNAm and target gene expression. **D** In samples with low DNA methylation, target gene expression is observed to be repressed as TF activity increased. On the other hand, in samples with high DNA methylation, target gene expression is relatively independent of TF activity. Therefore, DNA methylation is predicted to attenuate TF activity on the target gene. **DNAm**: DNA methylation

**A**

| | |
|---|---|
| Genome of reference | hg19 |
| Region ID | chr20:36148791−36148792 |
| Probe ID | cg17643025 |
| Target gene ID | ENSG00000053438 |
| Target gene Symbol | NNAT |
| TF gene ID | ENSG00000072310 |
| TF gene Symbol | SREBF1 |
| TF role | Activator |
| DNAm effect | Attenuating |

| Target ~ TF + DNAm Quant. Group + TF * DNAm Quant. Group | Estimate | P−Values |
|---|---|---|
| Direct effect of DNAm | 2.13 | 0.001903 |
| Direct effect of TF | 5.49 | 0.001054 |
| Synergistic effect of DNAm and TF | −6.86 | 0.001722 |

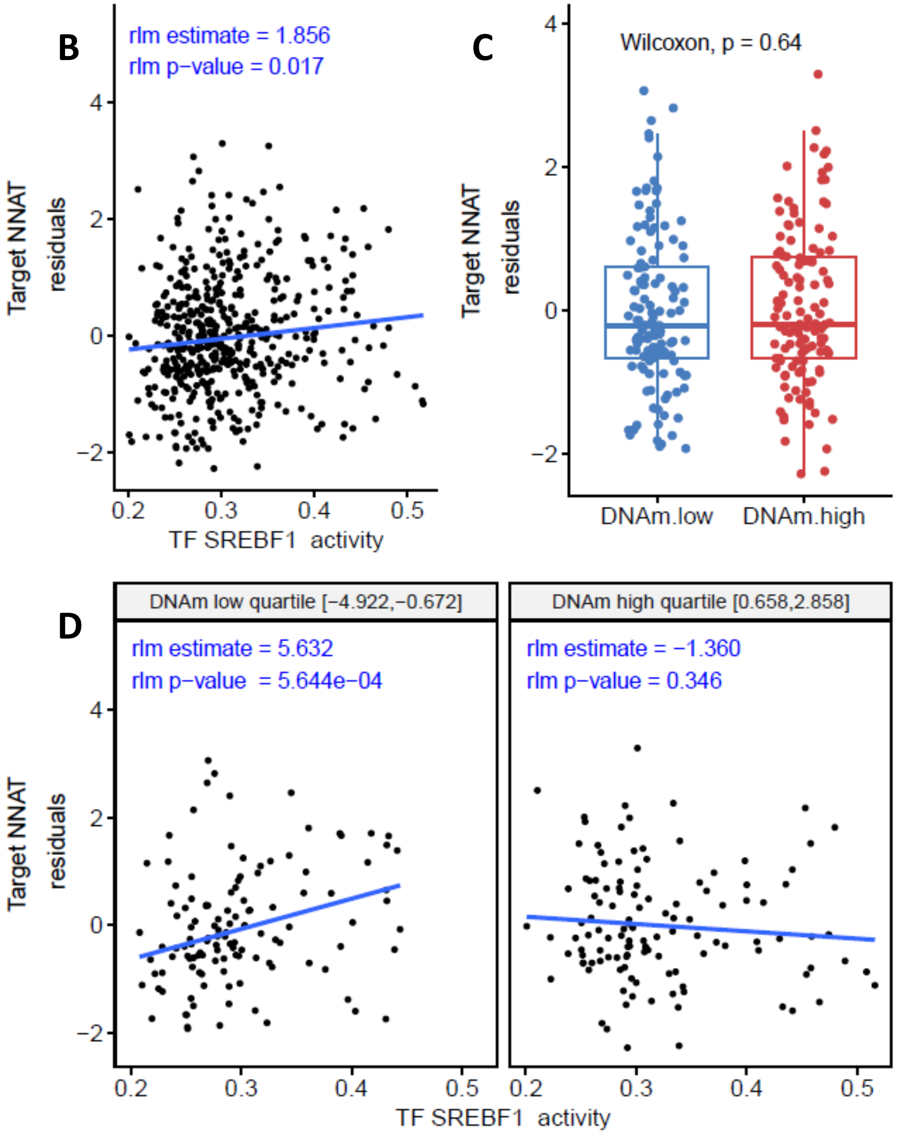


**Supplementary Figure 6** Results from MethReg analysis of ADNI blood samples dataset showed for the cg17643025-SREBF1-NNAT triplet, DNAm attenuated TF activity. **A** Meta data for the CpG-TF-target gene triplet and results of fitting robust linear model to the triplet dataset. **B** When all samples are considered, increased TF activity is associated with higher target gene expression levels. **C** Comparison of target gene expression between the DNAm groups showed no association between DNAm and target gene expression. **D** In samples with low methylation, target gene expression is observed to be increased as TF activity increased. In samples with high methylation, target gene expression is relatively independent of TF activity. Therefore, DNA methylation at cg17643025 is predicted to attenuate TF activity. **DNAm**: DNA methylation

**A**

| | |
|---|---|
| Genome of reference | hg19 |
| Region ID | chr17:80279310−80279311 |
| Probe ID | cg26312191 |
| Target gene ID | ENSG00000141574 |
| Target gene Symbol | SECTM1 |
| TF gene ID | ENSG00000196092 |
| TF gene Symbol | PAX5 |
| TF role | Activator |
| DNAm effect | Attenuating |

| Target ~ TF + DNAm Quant. Group + TF * DNAm Quant. Group | Estimate | P−Values |
|---|---|---|
| Direct effect of DNAm | 0.0161 | 0.920453 |
| Direct effect of TF | 0.281 | 1.45e−05 |
| Synergistic effect of DNAm and TF | −0.421 | 1.97e−05 |

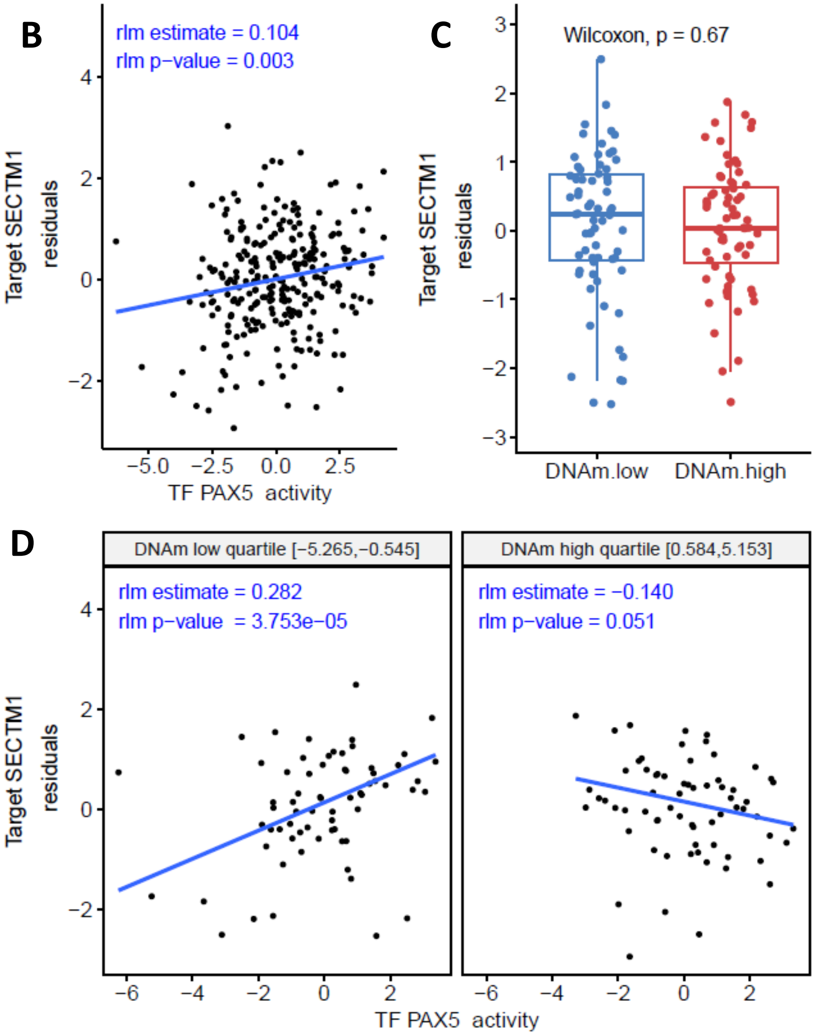


**Supplementary Figure 7** Results from MethReg analysis of ADNI blood samples dataset showed for the cg26312191-PAX5-SECTM1 triplet, DNAm attenuated TF activity. **A** Meta data for the CpG-TF-target gene triplet and results of fitting robust linear model to the triplet dataset. **B** When all samples are considered, increased TF activity is associated with higher target gene expression levels. **C** Comparison of target gene expression between the DNAm groups showed no association between DNAm and target gene expression. **D** In samples with low methylation, target gene expression is observed to be increased as TF activity increased. In samples with high methylation, target gene expression is relatively independent of TF activity. Therefore, DNA methylation at cg26312191 is predicted to attenuate TF activity. **DNAm**: DNA methylation

**A**

| | |
|---|---|
| Genome of reference | hg19 |
| Region ID | chr11:47376754−47376755 |
| Probe ID | cg01539849 |
| Target gene ID | ENSG00000110514 |
| Target gene Symbol | MADD |
| TF gene ID | ENSG00000185022 |
| TF gene Symbol | MAFF |
| TF role | Repressor |
| DNAm effect | Enhancing |

| Target ~ TF + DNAm Quant. Group + TF * DNAm Quant. Group | Estimate | P−Values |
|---|---|---|
| Direct effect of DNAm | 1.73 | 0.000166 |
| Direct effect of TF | 2.33 | 0.027182 |
| Synergistic effect of DNAm and TF | −6.57 | 5.22e−06 |

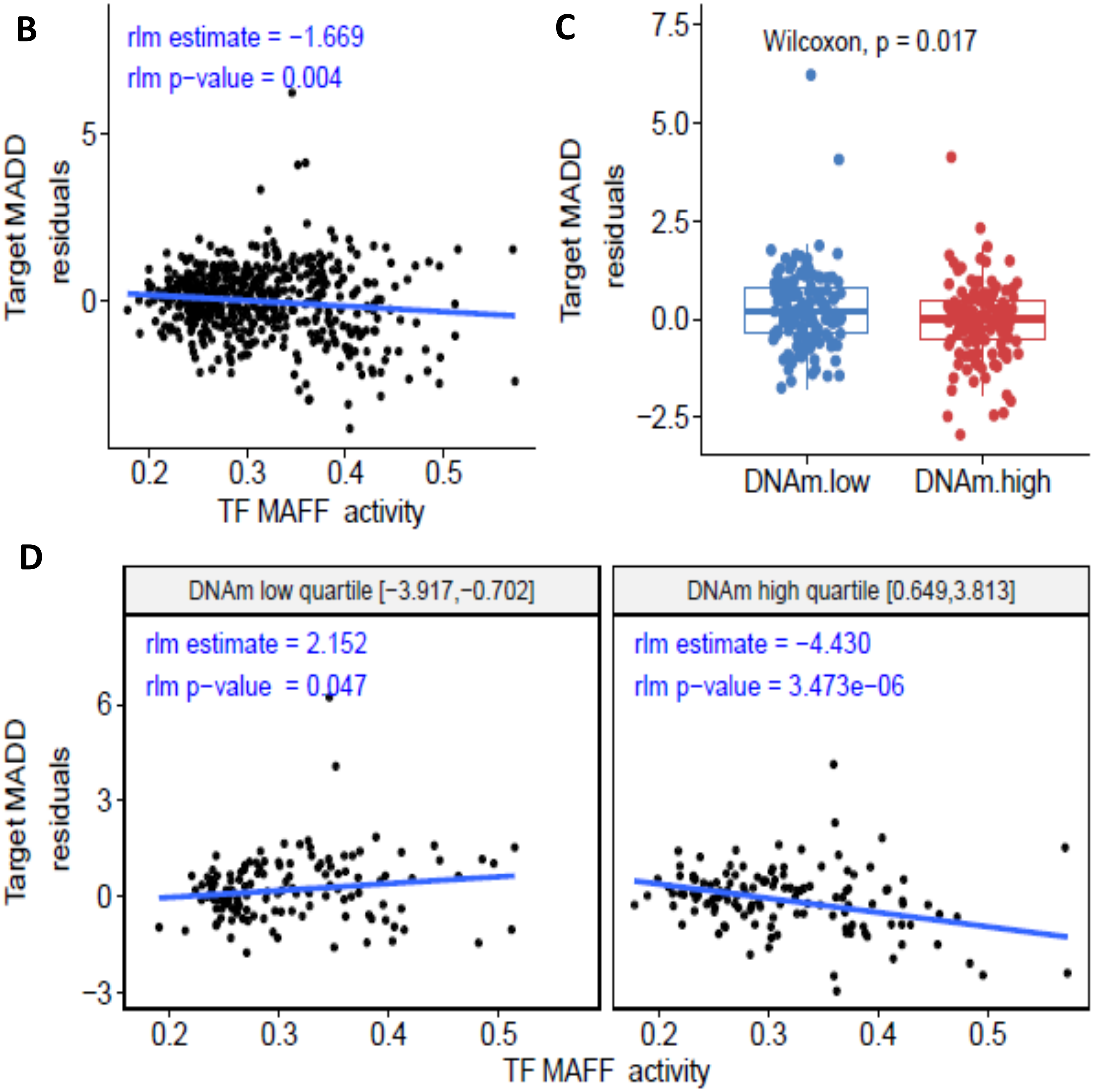


**Supplementary Figure 8** Results from MethReg analysis of ROSMAP brain samples dataset showed for the cg01539849-MAFF-MADD triplet, DNAm enhanced TF activity. **A** Meta data for the CpG-TF-target gene triplet and results of fitting robust linear model to the triplet dataset. **B** When all samples are considered, increased TF activity is associated with lower target gene expression levels. **C** Comparison of target gene expression between the DNAm groups showed higher target gene expression in samples with lower DNAm. **D** In samples with low methylation, target gene expression is relatively independent of TF activity. In samples with high methylation, target gene expression is observed to be decreased as TF activity increased. Therefore, DNA methylation at cg01539849-MAFF-MADD is predicted to enhance TF activity. **DNAm**: DNA methylation

**A**

| | |
|---|---|
| Genome of reference | hg19 |
| Region ID | chr19:8590567-8590568 |
| Probe ID | cg22568423 |
| Target gene ID | ENSG00000133246 |
| Target gene Symbol | PRAM1 |
| TF gene ID | ENSG00000170365 |
| TF gene Symbol | SMAD1 |
| TF role | Repressor |
| DNAm effect | Attenuating |

| Target ~ TF + DNAm Quant. Group + TF * DNAm Quant. Group | Estimate | P−Values |
|---|---|---|
| Direct effect of DNAm | −2.24 | 0.000320 |
| Direct effect of TF | −4.62 | 0.001031 |
| Synergistic effect of DNAm and TF | 8.71 | 1.60e−05 |

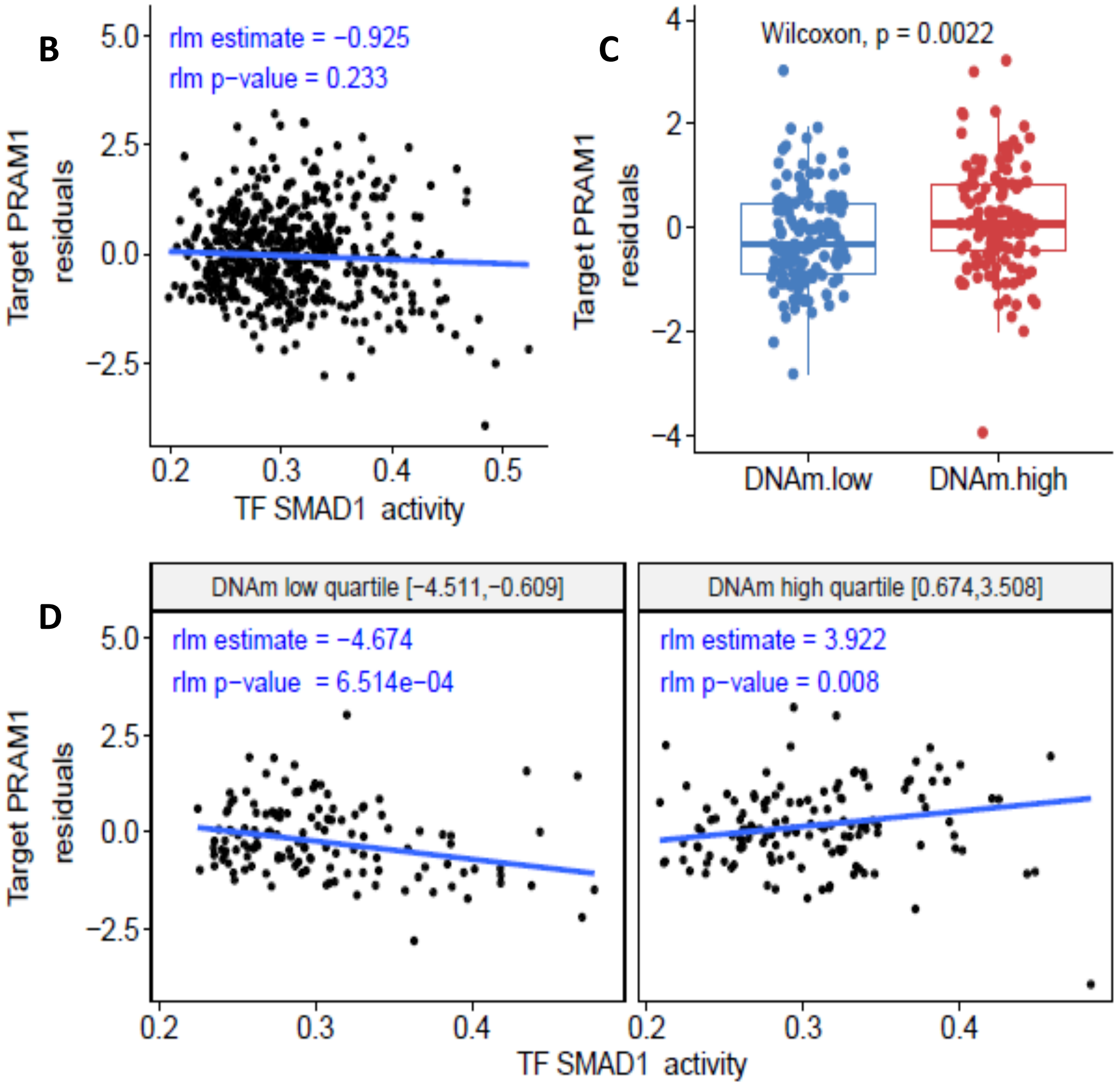


**Supplementary Figure 9** Results from MethReg analysis of ROSMAP brain samples dataset showed for the cg22568423-PRAM1-SMAD1, DNAm attenuated TF activity. **A** Meta data for the CpG-TF-target gene triplet and results of fitting robust linear model to the triplet dataset. **B** When all samples are considered, TF activity is not associated with target gene expression levels. **C** Comparison of target gene expression between the DNAm groups showed lower target gene expressions in samples with lower DNAm. **D** In samples with low methylation, target gene expression is observed to be decreased as TF activity increased. In samples with high methylation, the TF-target gene association is weaker. Therefore, DNA methylation at cg22568423 is predicted to attenuate TF activity. **DNAm**: DNA methylation

**A**

| Genome of reference | hg19 |
|---|---|
| Region ID | chr8:94924611−94924612 |
| Probe ID | cg02459543 |
| Target gene ID | ENSG00000164953 |
| Target gene Symbol | TMEM67 |
| TF gene ID | ENSG00000245848 |
| TF gene Symbol | CEBPA |
| TF role | Activator |
| DNAm effect | Enhancing |

| Target ~ TF + DNAm Quant. Group + TF * DNAm Quant. Group | Estimate | P−Values |
|---|---|---|
| Direct effect of DNAm | −1.83 | 0.000398 |
| Direct effect of TF | −3.5 | 0.002819 |
| Synergistic effect of DNAm and TF | 7.2 | 1.67e−05 |

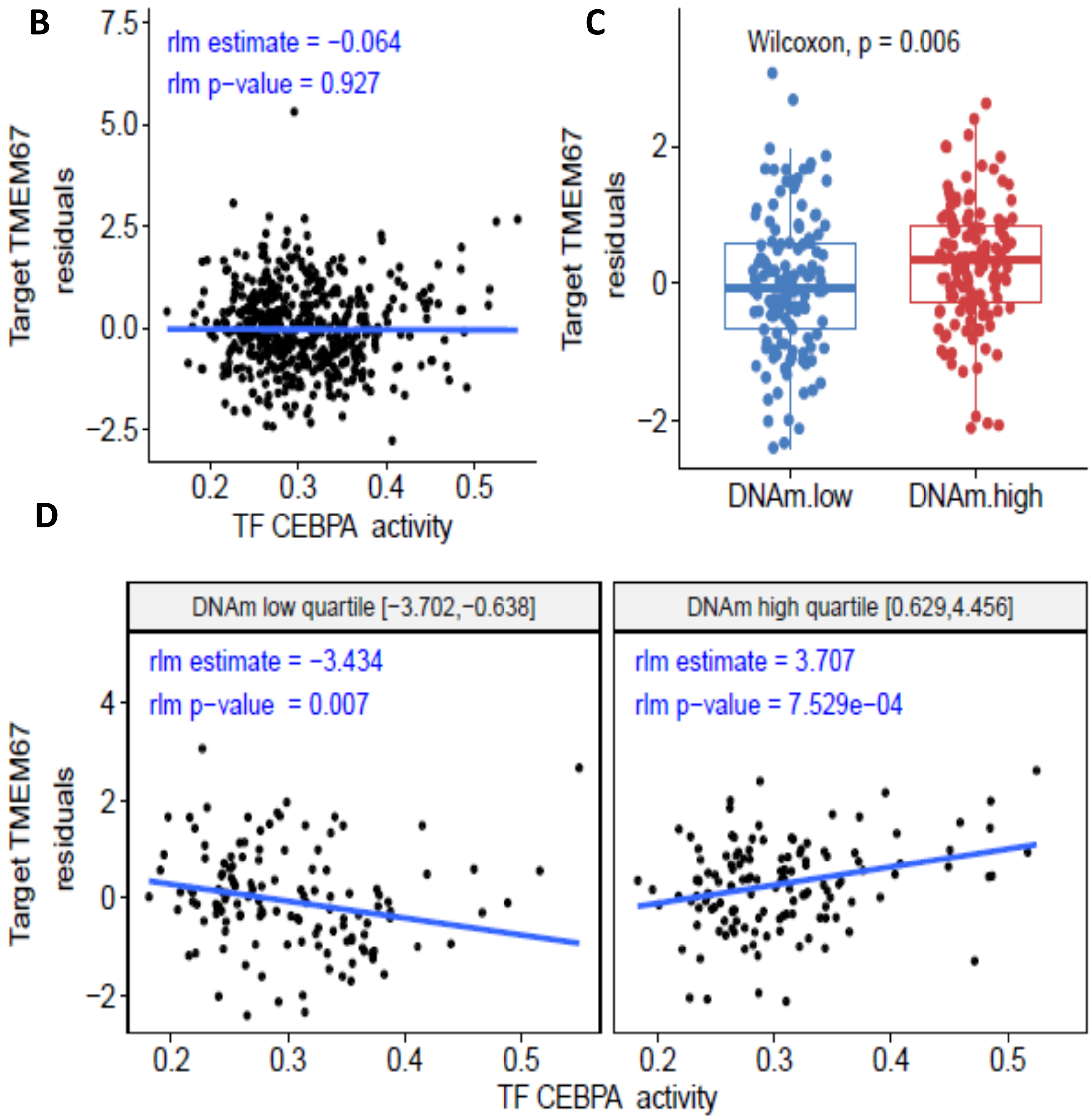


**Supplementary Figure 10** Results from MethReg analysis of ROSMAP brain samples dataset showed for the cg02459543-TMEM67-CEBPA triplet, DNAm enhanced TF activity. **A** Meta data for the CpG-TF-target gene triplet and results of fitting robust linear model to the triplet dataset. **B** When all samples are considered, TF activity is not associated with target gene expression levels. **C** Comparison of target gene expression between the DNAm groups showed lower target gene expressions in samples with lower DNAm. **D** In samples with high methylation, target gene expression is observed to be increased as TF activity increased. In samples with low methylation, TF-target gene expression is relatively weaker. Therefore, DNAm at cg02459543M is predicted to enhance TF activity. **DNAm:** DNA methylation

**A**

| | |
|---|---|
| Genome of reference | hg19 |
| Region ID | chr19:8590567-8590568 |
| Probe ID | cg22568423 |
| Target gene ID | ENSG00000142347 |
| Target gene Symbol | MYO1F |
| TF gene ID | ENSG00000245848 |
| TF gene Symbol | CEBPA |
| TF role | Repressor |
| DNAm effect | Attenuating |

| Target ~ TF + DNAm Quant. Group + TF * DNAm Quant. Group | Estimate | P-Values |
|---|---|---|
| Direct effect of DNAm | −1.56 | 0.002608 |
| Direct effect of TF | −4.12 | 5.45e−05 |
| Synergistic effect of DNAm and TF | 6.88 | 5.09e−05 |

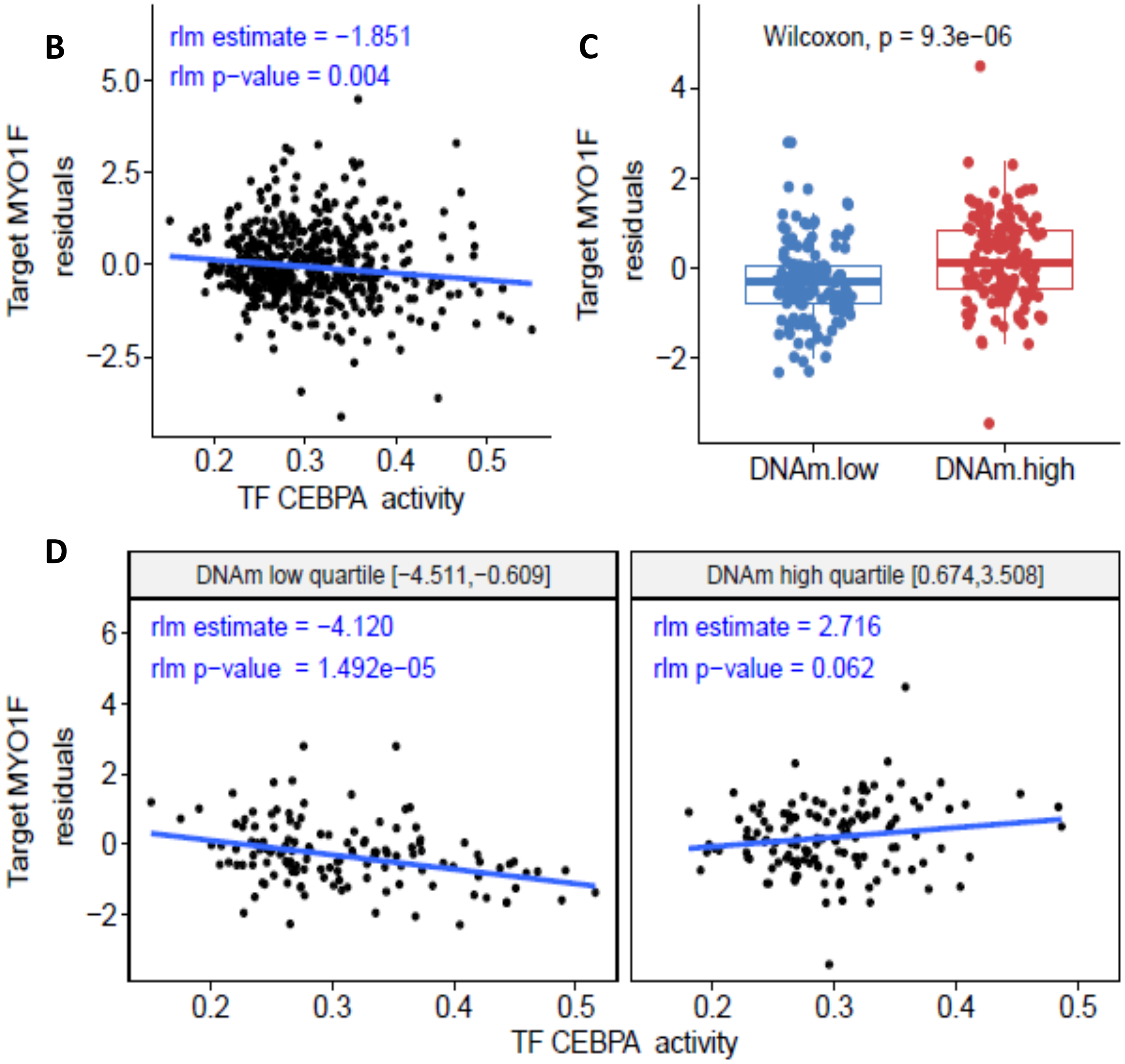


**Supplementary Figure 11** Results from MethReg analysis of ROSMAP brain samples dataset showed for the cg22568423-MYO1F-CEBPA, DNAm attenuated TF activity. **A** Meta data for the CpG-TF-target gene triplet and results of fitting robust linear model to the triplet dataset. **B** When all samples are considered, increased TF activity is associated with lower target gene expression levels. **C** Comparison of target gene expression between the DNAm groups showed lower target gene expressions in samples with lower DNAm. **D** In samples with low methylation, target gene expression is observed to be decreased as TF activity increased. In samples with high methylation, target gene expression is relatively independent of TF activity. Therefore, DNA methylation at cg22568423 is predicted to attenuate TF activity. **DNAm:** DNA methylation

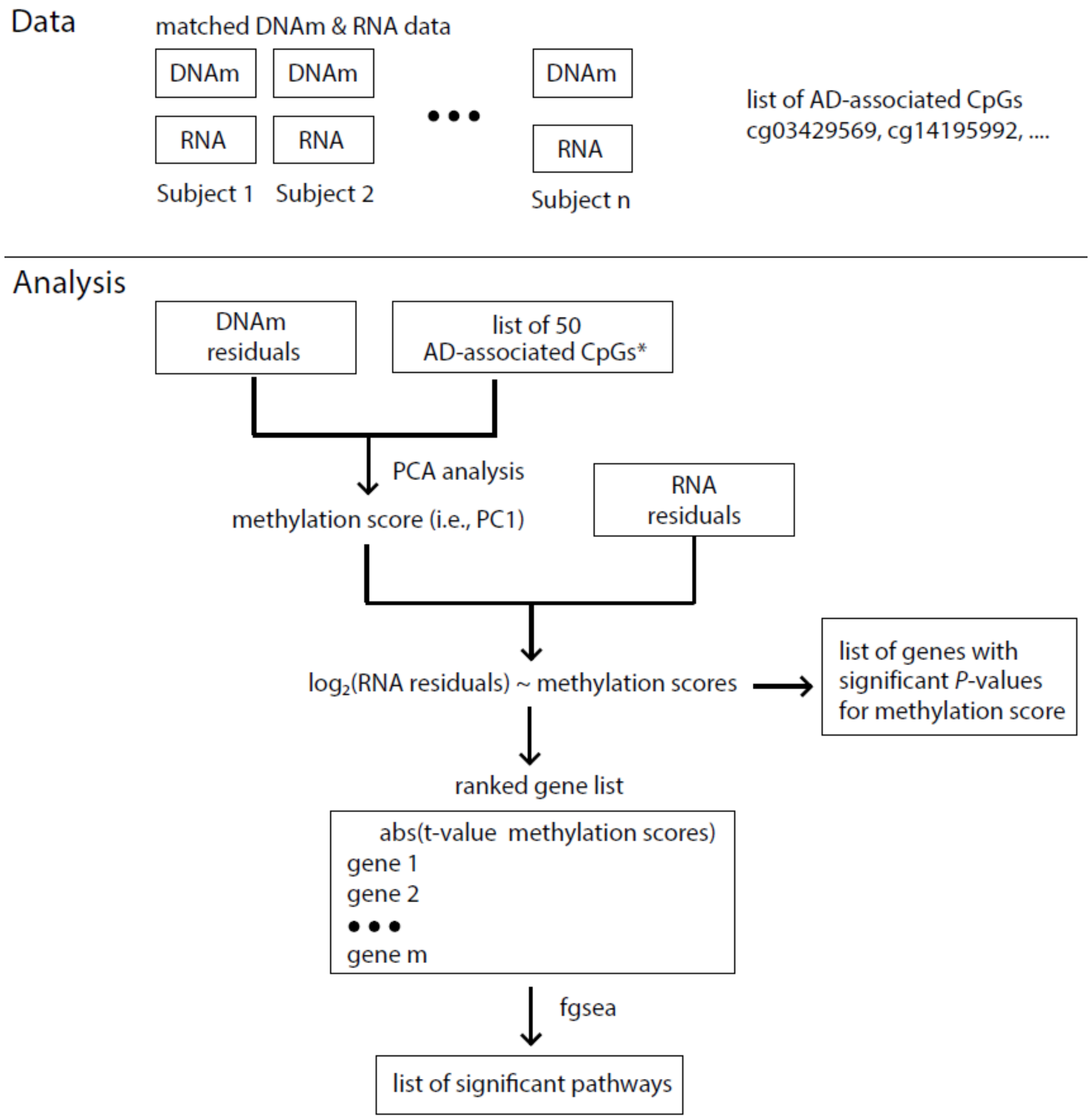


**Supplementary Figure 12** Workflow of integrative DNA methylation (DNAm) and gene expression datasets. For blood samples analysis, we used the ADNI dataset with 265 matched DNAm and gene expression samples. *DNAm residuals* were computed by fitting a linear model with methylation *M*-value as the outcome, age, sex, estimated immune cell-type proportions, and batch effects as independent variables and extracting residuals. *RNA residuals* were computed similarly, except by using log-transformed gene expression values as the outcome. For brain samples analysis, we used the ROSMAP AD dataset with 529 matched DNAm and RNA-seq samples. DNAm residuals and RNA residuals were computed in the same way as for blood samples, except that we used estimated neuron proportions in the DNAm model and expression levels of marker genes for different brain cell types in the RNA model (i.e., *ENO2* for neurons, *GFAP* for astrocytes, *CD68* for microglia, *OLIG2* for oligodendrocytes, and *CD34* for endothelial cells). For each tissue, we next performed the following analysis steps: (1) first, we used principal component analysis to summarize DNAm residuals at the significant CpGs by the first PC (PC1), which is a weighted linear combination of the methylation residuals, these are the methylation PC scores; (2) Next, we tested the association between methylation PC scores (i.e., PC1) and genome-wide gene expressions residuals using linear models; (3) Finally, we ranked the genes by the absolute value of t-statistics for methylation PC scores, and performed pathway analysis using the fgsea software.

* In blood samples analysis, we computed PC1 of the 50 AD-associated CpGs with $P < 10^{-5}$. In brain samples analysis, we computed PC1 of the 3751 FDR significant CpGs in our previous brain samples meta-analysis (Zhang et al. 2020 PMID: 33257653).
